# Adherence trajectory as an on-treatment risk indicator among drug-resistant TB patients in the Philippines

**DOI:** 10.1101/2022.05.04.22274664

**Authors:** S Huddart, DM Geocaniga-Gaviola, R Crowder, AR Lim, E Lopez, CL Valdez, CA Berger, Raul Destura, M Kato-Maeda, A Cattamanchi, AMC Garfin

## Abstract

**Introduction:** High levels of treatment adherence are critical for achieving optimal treatment outcomes among patients with tuberculosis (TB), especially for drug-resistant TB (DR TB). Current tools for identifying high-risk non-adherence are insufficient. Here, we apply trajectory analysis to characterize adherence behavior early in DR TB treatment and assess whether these patterns predict treatment outcomes.

**Methods:** We conducted a retrospective analysis of Philippines DR TB patients treated between 2013 and 2016. To identify unique patterns of adherence, we performed group-based trajectory modelling on adherence to the first 12 weeks of treatment. We estimated the association of adherence trajectory group with six-month and final treatment outcomes using univariable and multivariable logistic regression. We also estimated and compared the predictive accuracy of adherence trajectory group and a binary adherence threshold for treatment outcomes.

**Results:** Of 596 patients, 302 (50.7%) had multidrug resistant TB, 11 (1.8%) extremely drug-resistant (XDR) TB, and 283 (47.5%) pre-XDR TB. We identified three distinct adherence trajectories during the first 12 weeks of treatment: a high adherence group (n=483), a moderate adherence group (n=93) and a low adherence group (n=20). Similar patterns were identified at 4 and 8 weeks. Being in the 12-week moderate or low adherence group was associated with unfavorable six-month (adjusted OR [aOR] 3.42, 95% CI 1.90 - 6.12) and final (aOR 2.71, 95% 1.73 - 4.30) treatment outcomes. Adherence trajectory group performed similarly to a binary threshold classification for the prediction of final treatment outcomes (65.9 % vs. 65.4 % correctly classified), but was more accurate for prediction of six-month treatment outcomes (79.4% vs. 60.0% correctly classified).

**Conclusions:** Adherence patterns are strongly predictive of patient treatment outcomes. Trajectory-based analyses represent an exciting avenue of research into TB patient adherence behavior seeking to inform interventions which rapidly identify and support patients with high-risk adherence patterns.

## Introduction

High levels of treatment adherence are critical for achieving optimal treatment outcomes among patients with tuberculosis (TB). However, the length of TB treatment in addition to drug side effects can complicate adherence for patients. Adherence can also be influenced by elements of patients’ day-to-day lives including type of employment and proximity to a TB treatment center.^1^ Consequently, many patients miss doses or are lost to follow-up prior to treatment completion.

Current tools for identifying and ameliorating high-risk non-adherence are insufficient. Most TB programs rely on heuristic measures for flagging patients who may benefit from additional treatment support, extended TB treatment, or change in treatment regimen. There is little evidence to support the accuracy of these metrics which include missing a certain number of doses in a row or failing to take a certain number of doses by a given point in treatment. Adherence behavior, however, remains an appealing programmatic risk indicator. It is one of the few metrics which can be assessed while treatment is ongoing thus allowing for early intervention.

Advances in digital adherence technologies (DATs) make it increasingly feasible for TB programs to view and analyze day-by-day adherence data in real time. As many programs shift to all oral regimens, this rapid and data-driven responsiveness may become possible even for the lengthy regimens used to treat drug-resistant TB (DR TB). Thus, there is a critical need to understand how day-to-day adherence behaviors early in DR TB treatment influence treatment outcomes. Such information is critical to inform future interventions for patients demonstrating high-risk adherence patterns.

A large patient-level pooled analysis identified adherence below 90% as a key driver of treatment failure among patients with drug susceptible TB.^2^ Similarly, we recently showed that overall adherence proportion is equally important to the success of DR TB.^3^ These analyses summarized adherence as the proportion of doses taken out of all scheduled doses across the entire treatment period. However, condensing longitudinal adherence trends into a single measure assumes that the distribution of missed doses will not impact patient outcomes, and does not allow for assessment of how early adherence behavior may predict treatment outcomes.

Increasingly popular methods exist which can assess entire longitudinal trajectories and this trajectory-based approach has been shown to improve outcome prediction relative to single summary measures in TB^4^ and other health conditions.^5,6^ Here, we apply a trajectory-based analysis to characterize adherence behavior early in DR TB treatment and assess whether these patterns predict treatment outcomes.

## Methods

### Study setting

The Philippines is one of the WHO high-burden countries for overall TB burden as well as DR TB. Through the Philippines National TB Program (NTP), TB patients receive first-line drug susceptibility testing (DST) via a network of TB treatment units. All patients with detected isoniazid and rifampicin resistance receive second line DST with samples tested at the National Tuberculosis Reference Laboratory or other tertiary laboratories. DR TB patients receive standardized treatment regimens administered via Directly Observed Therapy (DOT) at DR TB treatment units.

### Study design and patient population

We conducted a retrospective analysis of patients enrolled in a parent cohort study evaluating targeted next-generation deep sequencing to characterize mutations associated with drug resistance. The parent cohort included patients with extensively drug-resistant (XDR) or pre-XDR TB and a random 1:1 sample of patients with multidrug-resistant (MDR) TB confirmed as DR TB through the Philippines National TB program between 2013 and 2016 (n=672).^3^ Some included patients may have initiated treatment outside of this time period depending on when they initiated treatment relative to final confirmation of DR TB status. For this analysis, we excluded patients who died or were lost to follow-up within the first 12 weeks of treatment initiation. Institutional review boards at the University of California San Francisco (IRB # 16-18610, Reference # 290984), the University of the Philippines Manila (UPMREB 2016-122-01), and the Research Institute for Tropical Medicine (2016-45) approved the study and provided a waiver of informed consent for patient-level data extraction and analysis.

### Data collection

Patient demographics and other baseline variables were collected from the Philippines Integrated Tuberculosis Information Services electronic database. At each DR TB treatment unit, patients’ daily dosing information and outcome data were recorded onto a paper treatment card. Study staff or NTP staff uploaded photos of the treatment cards of study patients to a secure server. Study staff then extracted patient demographic and clinical information into a secure Research Electronic Data Capture (REDCap) database using a standardized data extraction tool.^7^

### Definitions and outcomes

We defined patients as adherent for a given day if their treatment card recorded that they had taken medication on that day. We defined the treatment period for which we calculated adherence as the period from the day of treatment initiation to the day of treatment outcome. We additionally corrected daily adherence data for complete drug holds where patients (n=9) were instructed to stop all medications for a period of time due to adverse drug reactions. These patients were marked as adherent for the duration of the complete drug hold. Of these patients, only 3 had complete drug holds within the first 12 weeks of treatment. For final treatment outcomes, favorable treatment outcome was defined as either cure or completion of treatment. Unfavorable outcome was defined as treatment failure, death, or loss to follow-up. For six-month treatment outcomes, favorable outcome was defined as being retained in care and unfavorable outcome was defined as death or loss to follow-up. For the majority of the study period, the NTP program used a 6 day a week dosing schedule. In 2017, some treatment units began to phase in a 7 day a week dosing regimen. For consistency, we have treated Sundays as a drug holiday for all patients for the duration of treatment.

Our primary objective was to estimate the association between adherence patterns observed in the first 12 weeks of treatment and final treatment outcome. Our secondary objectives were to estimate the association between adherence patterns and six-month treatment outcomes as well as to characterize the predictive accuracy of adherence patterns for six-month and final treatment outcomes.

### Data analysis

To identify and characterize unique patterns of adherence, we performed group-based trajectory modelling, a type of finite mixture modeling that simultaneously estimates group membership and group mean trajectory.^8,9^ Specifically, we constructed and modeled weekly adherence trajectories (the sum of doses taken each week; weekly maximum of 6) over the first 12 weeks of treatment for each patient. We considered models with between two and five unique trajectory groups and selected the best model based on Akaike Information Criteria (AIC), Bayesian Information Criteria (BIC), and absolute fit to the data. We then repeated the modeling to estimate group membership using 4-week and 8-week adherence trajectories.

To determine the association of adherence trajectory group with final and six-month treatment outcome, we performed univariable and multivariable logistic regression. The multivariable model adjusted for patient clinical characteristics available in programmatic data: age, sex, category of drug resistance, sputum grade, region, year of treatment, chest X-ray status (cavitary, noncavitary, missing) and body mass index (BMI). For BMI, height was imputed when missing (n=132) using the median height for the respective sex.

Last, we estimated and compared the predictive accuracy of adherence trajectory group membership and a binary adherence threshold (>=90%) for outcomes at six months and the end of treatment. We selected a 90% binary adherence threshold based on literature demonstrating that this was a critical value for successful treatment of drug-susceptible TB.^2^ We calculated the sensitivity, specificity and proportion of patients correctly classified by adherence trajectory group membership based on 4-, 8- and 12-weeks of initial treatment adherence data as a predictor of final treatment outcome and six-month treatment outcome. We calculated the same predictive performance metrics for an adherence trajectory group model which additionally included baseline patient characteristics and a model using the binary threshold.

## Results

### Patient characteristics

Of 672 patients with drug-resistant TB included in the parent cohort, 76 (11.3%) experienced a treatment outcome within the first 12 weeks of treatment and were excluded. The remaining 596 patients (Table 1) included 302 (50.7%) with MDR, 283 (47.5%) with pre-XDR, and 11 (1.8%) with XDR TB. Median patient age was 41 years (IQR 29-51), 182 (30.5%) patients were female and the median BMI at treatment initiation was 18.1 (IQR 16.0-21.1). Median adherence was 78.7% (IQR 61.3-92.7%) over the full course of treatment and 91.7% (IQR 76.4-97.2%) during the first 12 weeks of treatment. 222 patients (37.2%) patients had an unfavorable treatment outcome.

**Table 1.** Cohort demographics by adherence pattern at 12 weeks

|  | High (n=483) | Moderate (n=93) | Low (n=20) | P-value | Overall (n=596) |
| --- | --- | --- | --- | --- | --- |
| **Age** |  |  |  |  |  |
| Median [Min, Max] | 40.0 [14.0, 76.0] | 41.0 [19.0, 75.0] | 42.0 [22.0, 64.0] | 0.39 | 41.0 [14.0, 76.0] |
| **Sex** |  |  |  |  |  |
| Male | 339 (70.2%) | 61 (65.6%) | 14 (70.0%) | 0.68 | 414 (69.5%) |
| Female | 144 (29.8%) | 32 (34.4%) | 6 (30.0%) |  | 182 (30.5%) |
| **Resistance category** |  |  |  |  |  |
| MDR | 249 (51.6%) | 45 (48.4%) | 8 (40.0%) | 0.64 | 302 (50.7%) |
| Pre-XDR | 225 (46.6%) | 47 (50.5%) | 11 (55.0%) |  | 283 (47.5%) |
| XDR | 9 (1.9%) | 1 (1.1%) | 1 (5.0%) |  | 11 (1.8%) |
| **Year of initiation** |  |  |  |  |  |
| 2012 | 9 (1.9%) | 1 (1.1%) | 0 (0%) | 0.53 | 10 (1.7%) |
| 2013 | 47 (9.7%) | 9 (9.7%) | 4 (20.0%) |  | 60 (10.1%) |
| 2014 | 128 (26.5%) | 17 (18.3%) | 3 (15.0%) |  | 148 (24.8%) |
| 2015 | 206 (42.7%) | 46 (49.5%) | 8 (40.0%) |  | 260 (43.6%) |
| 2016/17 | 93 (19.3%) | 20 (21.5%) | 5 (25.0%) |  | 118 (19.8%) |
| **Region** |  |  |  |  |  |
| Ilocos Region | 2 (0.4%) | 1 (1.1%) | 0 (0%) | 0.88 | 3 (0.5%) |
| Cagayan Valley | 43 (8.9%) | 4 (4.3%) | 2 (10.0%) |  | 49 (8.2%) |
| Central Luzon | 5 (1.0%) | 1 (1.1%) | 0 (0%) |  | 6 (1.0%) |
| Calabarzon | 93 (19.3%) | 17 (18.3%) | 4 (20.0%) |  | 114 (19.1%) |
| Mimaropa | 13 (2.7%) | 3 (3.2%) | 1 (5.0%) |  | 17 (2.9%) |
| Bicol Region | 28 (5.8%) | 11 (11.8%) | 1 (5.0%) |  | 40 (6.7%) |
| Northern Mindanao | 6 (1.2%) | 0 (0%) | 0 (0%) |  | 6 (1.0%) |
| Davao Region | 4 (0.8%) | 0 (0%) | 0 (0%) |  | 4 (0.7%) |
| Soccsksargen | 36 (7.5%) | 8 (8.6%) | 0 (0%) |  | 44 (7.4%) |
| Caraga | 9 (1.9%) | 3 (3.2%) | 0 (0%) |  | 12 (2.0%) |
| Zamboanga Peninsula | 192 (39.8%) | 34 (36.6%) | 11 (55.0%) |  | 237 (39.8%) |
| Autonomous Region in Muslim Mindanao | 32 (6.6%) | 5 (5.4%) | 1 (5.0%) |  | 38 (6.4%) |
| Cordillera Administrative Region | 12 (2.5%) | 5 (5.4%) | 0 (0%) |  | 17 (2.9%) |
| National Capital Region | 8 (1.7%) | 1 (1.1%) | 0 (0%) |  | 9 (1.5%) |
| **CXR cavitation** |  |  |  |  |  |
| No | 162 (33.5%) | 41 (44.1%) | 12 (60.0%) | 0.04 | 215 (36.1%) |
| Yes | 187 (38.7%) | 27 (29.0%) | 6 (30.0%) |  | 220 (36.9%) |
| Missing | 134 (27.7%) | 25 (26.9%) | 2 (10.0%) |  | 161 (27.0%) |
| **Sputum Grade** |  |  |  |  |  |
| Negative (0) | 74 (15.3%) | 18 (19.4%) | 3 (15.0%) | 0.77 | 95 (15.9%) |
| n+ | 61 (12.6%) | 9 (9.7%) | 0 (0%) |  | 70 (11.7%) |
| 1+ | 125 (25.9%) | 23 (24.7%) | 5 (25.0%) |  | 153 (25.7%) |
| 2+ | 100 (20.7%) | 21 (22.6%) | 5 (25.0%) |  | 126 (21.1%) |
| 3+ | 120 (24.8%) | 20 (21.5%) | 6 (30.0%) |  | 146 (24.5%) |
| Missing | 3 (0.6%) | 2 (2.2%) | 1 (5.0%) |  | 6 (1.0%) |
| **BMI** |  |  |  |  |  |
| Median [Min, Max] | 18.3 [9.90, 34.2] | 17.8 [11.2, 25.1] | 16.7 [11.7, 23.3] | 0.05 | 18.1 [9.90, 34.2] |
| **Overall adherence proportion** |  |  |  |  |  |
| Median [Min, Max] | 83.3% [25.7%, 100%] | 51.3% [17.3%, 92.9%] | 43.4 [9.1%, 85.1%] | **0.00** | 78.7% [9.1%, 100%] |
| **12 week adherence proportion** |  |  |  |  |  |
| Median [Min, Max] | 93.1 [65.3%, 100%] | 55.6% [18.1%, 76.4%] | 39.6% [15.3%, 61.1%] | **0.00** | 91.7% [15.3%, 100%] |
| **Treatment outcome** |  |  |  |  |  |
| Favorable | 327 (67.7%) | 45 (48.4%) | 2 (10.0%) | **0.00** | 374 (62.8%) |
| Unfavorable | 156 (32.3%) | 48 (51.6%) | 18 (90.0%) |  | 222 (37.2%) |

CXR- chest x-ray.
For the statistical tests of the distribution of variables across the adherence groups, one-way ANOVA was used for continuous variables and $\chi^2$ test for categorical variables.

### Adherence trajectories

When comparing models with between two and five unique adherence trajectory groups over the first 12 weeks of treatment, the three and four group trajectory models provided similar AIC and BIC metrics (S T1), but the three group model demonstrated the best absolute fit to the data (S F1). Using this model, three distinct adherence trajectories were identified during the first 12 weeks of treatment: a high adherence group (n=483), a moderate adherence group (n=93) and a low adherence group (n=20) (Figure 1). Membership in these adherence groups did not have a strong relationship to patient demographics (Table 1) except for CXR cavitation (p=0.04). Median adherence was 83.3%, 51.3%, and 43.4% over the full course of treatment and 93.1%, 55.6%, and 39.6% during the first 12 weeks for the high, moderate and low adherence groups, respectively.

**Figure 1.**
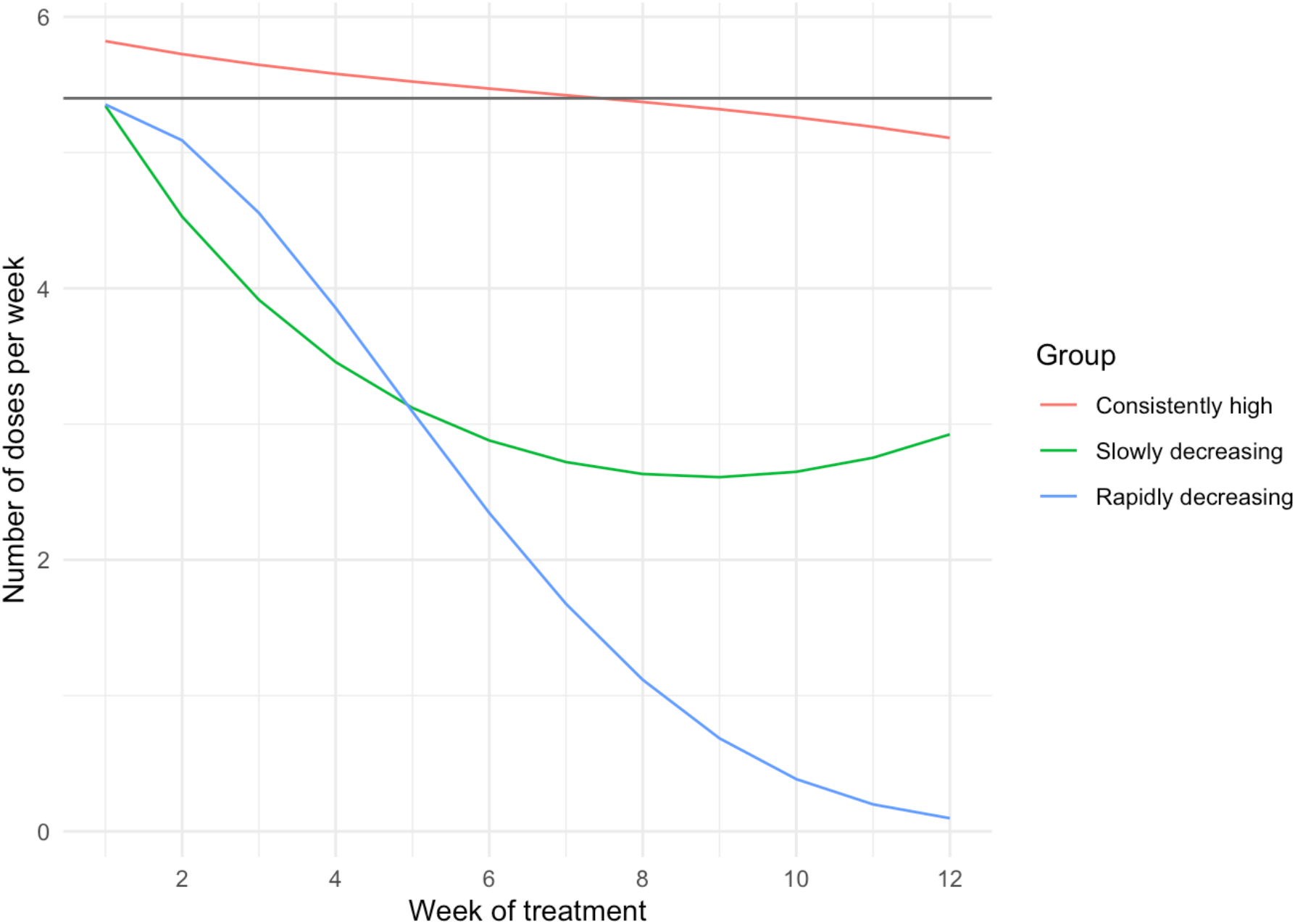
Adherence patterns identified by group-based trajectory modelling at 12 weeks. Dark grey horizontal line indicates 90% adherence threshold.

Distinct adherence patterns could be identified as early as four weeks into treatment (Figure 2). For these earlier timepoints, a two group model offered the best fit (S T1). The two adherence groups identified at four weeks and eight weeks follow a similar shape to the high and low adherence trajectory groups identified at 12 weeks.

**Figure 2.**
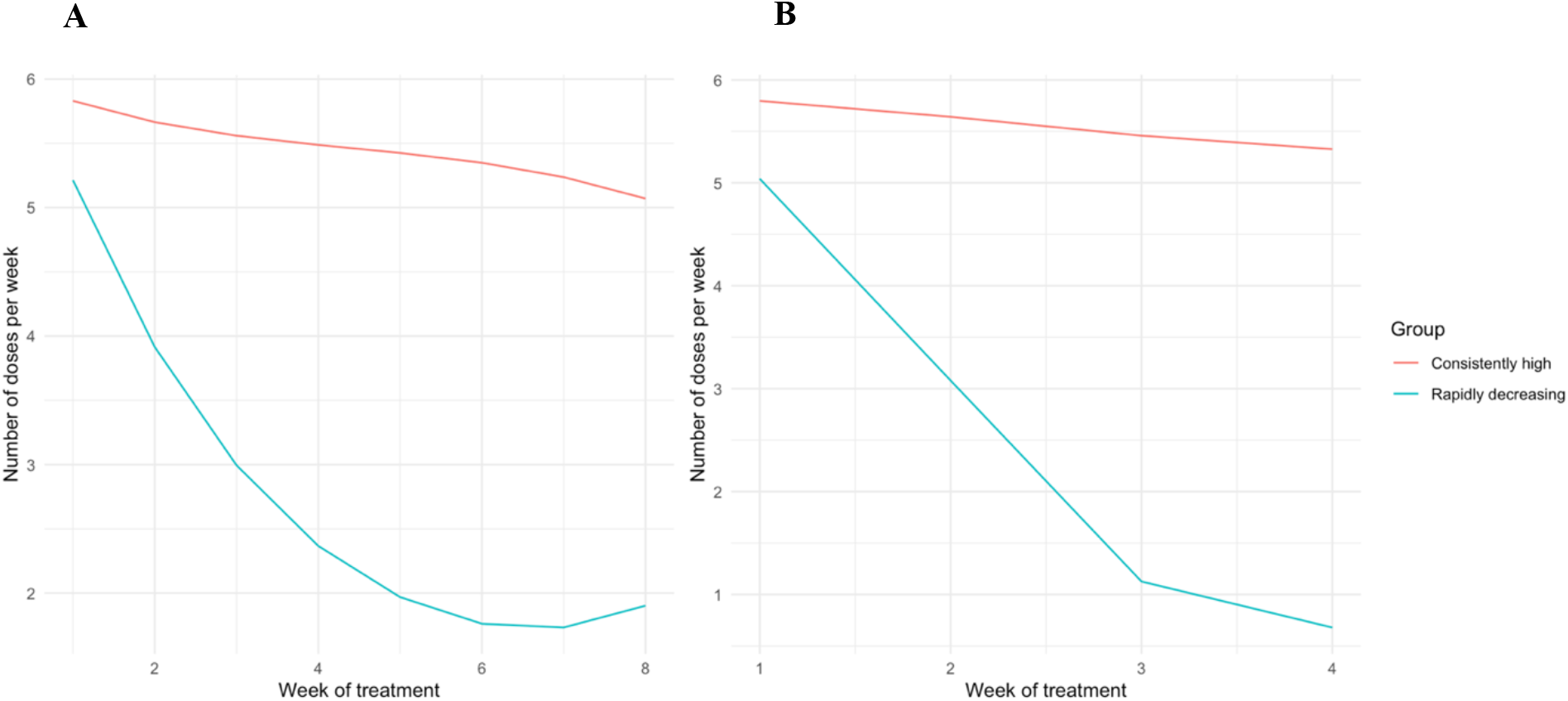
Adherence patterns identified by group-based trajectory modelling at (A) 8 weeks and (B) 4 weeks

### Association with final treatment outcome

Being in the moderate or low adherence group at 12 weeks was associated with significantly higher odds of unfavorable final treatment outcome in unadjusted (OR 2.94, 95% CI 1.94 - 4.47) and adjusted (OR 2.71, 95% 1.73 - 4.30) analyses (Table 2). The adjusted model was a significantly better fit than the model with only adherence trajectory groups (likelihood ratio test, p= 0.04).

**Table 2.** Coefficients from logistic regression model predicting final unfavorable treatment outcome based on 12 week adherence pattern, n=596

|  | OR (95% CI) |  |
| --- | --- | --- |
|  | Unadjusted | Adjusted |
| High adherence pattern | Reference | Reference |
| Moderate + Low adherence pattern | **2.94 (1.94, 4.47)** | **2.71 (1.73, 4.30)** |

*The adjusted model included patient sex, resistance category (MDR/pre-XDR/XDR), year of treatment initiation, region, chest x-ray cavitation (no/yes/missing), sputum grade and BMI.*

The percentage of treatment outcomes classified correctly was similar across models based on adherence pattern only or adherence plus baseline patient characteristics (Table 3), ranging from 63.9 to 65.9% and 65.4 to 69.1%, respectively, for 4-, 8- and 12-week adherence trajectory groups (Table 3). When using only adherence pattern, sensitivity was poor (range 6.3 – 29.7%) but specificity was high (range 87.4 to 98.1%). Sensitivity was higher (range 30.3 - 35.3%) when using adherence pattern plus baseline patient characteristics but specificity decreased (range 86.0 – 89.0%).

**Table 3.**
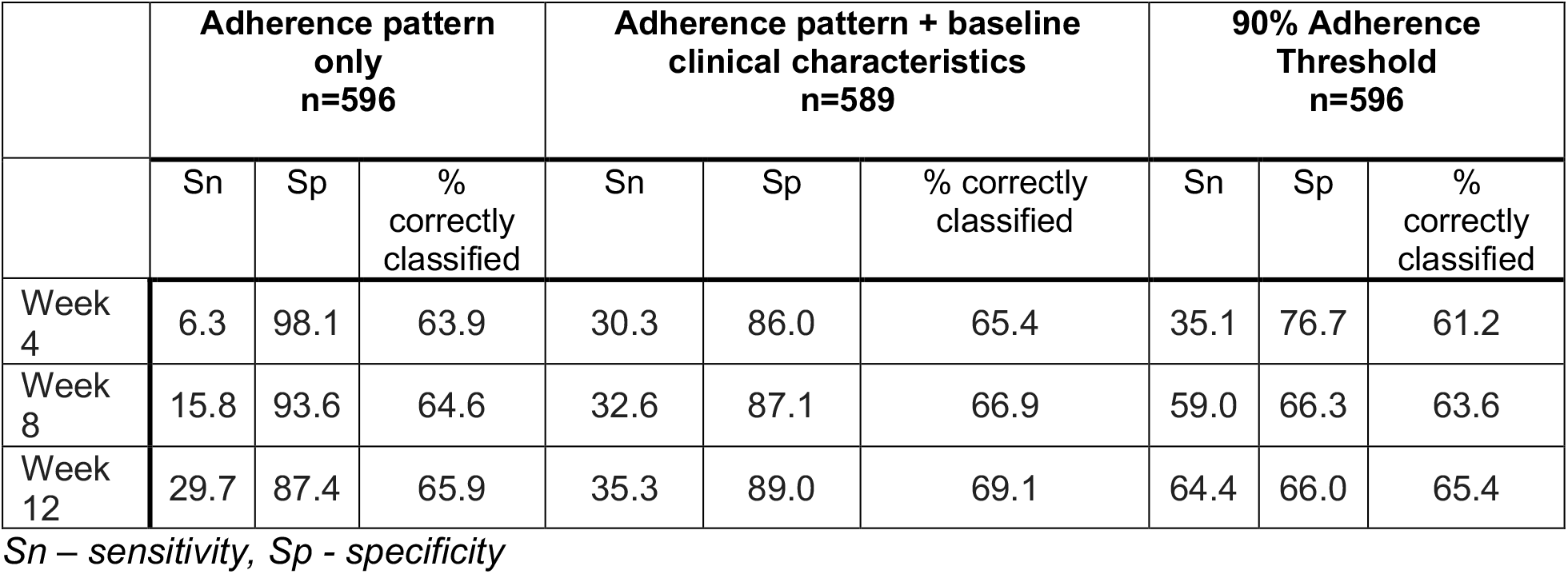
Accuracy of final treatment outcome prediction from 4 weeks to 12 weeks using only adherence pattern, adherence pattern and baseline clinical characteristics, and a 90% adherence threshold.

The predictive performance of trajectory groups identified using group-based trajectory modeling was no better than that using a simple binary threshold of 90% adherence to define low and high adherence groups (Table 3). The binary threshold correctly classified a similar percentage of treatment outcomes (range 61.2-65.4%). The binary threshold was more sensitive but less specific than the 12-week, 8-week or 4-week adherence pattern groups.

### Association with 6-month treatment outcome

Of 596 patients, 532 (89.3%) were retained in care at 6 months while 18 (3.0%) had died and 46 (7.7%) were lost to follow-up by this timepoint. Adherence pattern was significantly associated with 6-month treatment outcome in univariable analysis (OR 3.78, 95% CI 2.18 - 6.53) and maintained its significant association in multivariable analysis (OR 3.42, 95% CI 1.90 - 6.12, S T2).

The percentage of six-month outcomes classified correctly by a model using only adherence trajectory group was highest in the 4-week model (87.4%) and lowest in the 12-week model (79.4%, Table 4). Sensitivity of the adherence trajectory group model was poor (range 7.8 - 42.2%) and specificity was again higher (83.8 - 97.0%). The proportion of patients correctly classified by the model using adherence pattern and baseline clinical characteristics was higher (range 89.3 - 89.5%) than model using adherence pattern alone. Sensitivity for this model was lower (1.6 - 6.3%) and specificity was higher (99.8 - 100%) compared to the results of the model using only adherence pattern.

**Table 4.** Accuracy of 6-month outcome prediction from 4 weeks to 12 weeks using only adherence pattern, adherence pattern and baseline clinical characteristics, and a 90% adherence threshold.

|  | Adherence pattern only<br>n=596 |  |  | Adherence pattern + baseline clinical characteristics<br>n=589 |  |  | 90% Adherence Threshold<br>n=596 |  |  |
| --- | --- | --- | --- | --- | --- | --- | --- | --- | --- |
|  | Sn | Sp | % correctly classified | Sn | Sp | % correctly classified | Sn | Sp | % correctly classified |
| Week 4 | 7.8 | 97.0 | 87.4 | 1.6 | 100 | 89.3 | 34.4 | 73.1 | 69.0 |
| Week 8 | 25.0 | 92.0 | 84.7 | 6.3 | 99.8 | 89.6 | 62.5 | 59.2 | 60.0 |
| Week 12 | 42.2 | 83.8 | 79.4 | 4.7 | 99.8 | 89.5 | 75.0 | 58.3 | 60.0 |

*Sn – sensitivity, Sp - specificity*

The binary threshold model had worse performance for 6-month treatment outcomes than either of the adherence pattern models, correctly classifying 60.0 - 69.0% of patient outcomes. Sensitivity of the threshold classification ranged from 34.3 - 75.0% and specificity ranged from 58.3 – 73.1%.

## Discussion

In this work, we identified unique adherence patterns as early as four weeks into DR TB treatment. At 12 weeks into treatment, we identified three distinct treatment adherence patterns: consistently high, slowly decreasing and rapidly decreasing. Before 12 weeks, we identified two distinct patterns: consistently high and rapidly decreasing. These patterns were not strongly correlated with baseline patient data suggesting that they capture unique information about a patient. Further, these patterns were strongly associated with both 6-month and final treatment outcomes. Adherence pattern had moderate predictive capacity for treatment outcomes which could be slightly improved by adding patient clinical characteristics to the model. Adherence patterns did not outperform a simple binary threshold for prediction of outcomes at the end of treatment but did outperform the binary threshold for prediction of 6-month treatment outcome.

Adherence is a complex longitudinal behavior and we have demonstrated that multiple distinct patterns of this adherence behavior can exist within a single TB patient cohort. In line with other studies in this area, adherence was again strongly associated with treatment outcomes.^2^ While trajectory-based classifications did not always outperform binary threshold classifications when predicting treatment outcomes, trajectory-based analyses open up avenues of research into the drivers of distinct adherence patterns. As an example, among patients whose adherence fell below the 90% threshold, we identified two distinct patterns of nonadherence which may be caused by unique patient characteristics and experiences. These may range from the patient’s type of employment to the distance between a patient’s home and the treatment center, to the degree of social support each patient receives. Given the rise of DATs, real-time adherence data is increasingly available. Better understanding adherence patterns and their influence on treatment could allow for the development of algorithms to flag high-risk patients earlier and more accurately than the currently used heuristics.

This work contributes to evidence that adherence is a critical factor for successful TB treatment, especially during DR TB treatment. Our results are in line with other trajectory-based analyses of adherence in other conditions. Salmasi and colleagues found similarly complex adherence patterns within patients taking anticoagulants for atrial fibrillation.^10^ We found mixed results for the improved outcome prediction capacity of trajectory-based methods relative to binary classifications. This is in contrast with work by Liu and colleagues who demonstrated improved outcome prediction by using glycemic trajectories as a predictor rather than a simple diabetes diagnosis dichotomy.^4^

There are several limitations to this work. First, as this cohort represents programmatically available data, there are several clinical indicators of disease severity that were not available or had high degrees of missingness. These include smoking status as well as diagnoses of HIV, diabetes and liver disease. These indicators would likely have improved the accuracy of the models which included patient clinical characteristics. Further, we did not have data on patients’ socioeconomic status or other detailed demographics. This data may have revealed some of the factors driving the observed differences in adherence behavior. To our knowledge, this is the first trajectory-based analysis of TB patient adherence data. As such, we are not yet sure whether these patterns are generalizable to TB patients outside the Philippines or to patients being treated for drug-sensitive TB. Finally, only a small number of patients were classified in the rapidly decreasing adherence trajectory group which limited our power to detect unique characteristics about these patients.

In summary, trajectory-based analyses represent an exciting avenue of research into TB patient adherence behavior. Future studies which fully utilize adherence data, potentially collected by DATs, may lead to interventions which rapidly identify and support patients with high-risk adherence patterns.

## Data Availability

All data produced in the present study are available upon reasonable request to the authors with the approval of the overseeing IRB.

