## Supplemental Material for "Adherence trajectory as an on-treatment risk indicator among drug-resistant TB patients in the Philippines"

**S T1**, Cross tabulation of binary threshold and trajectory group

| | 4 weeks | | 8 weeks | | 12 weeks | |
|---|---|---|---|---|---|---|
| | AIC | BIC | AIC | BIC | AIC | BIC |
| 2 group model | **9070** | **9133** | **18303** | **18375** | 27561 | 27636 |
| 3 group model | 9116 | 9214 | 18466 | 18576 | **27476** | **27593** |
| 4 group model | No convergence | No convergence | No convergence | No convergence | **27397** | **27555** |
| 5 group model | No convergence | No convergence | No convergence | No convergence | No convergence | No convergence |

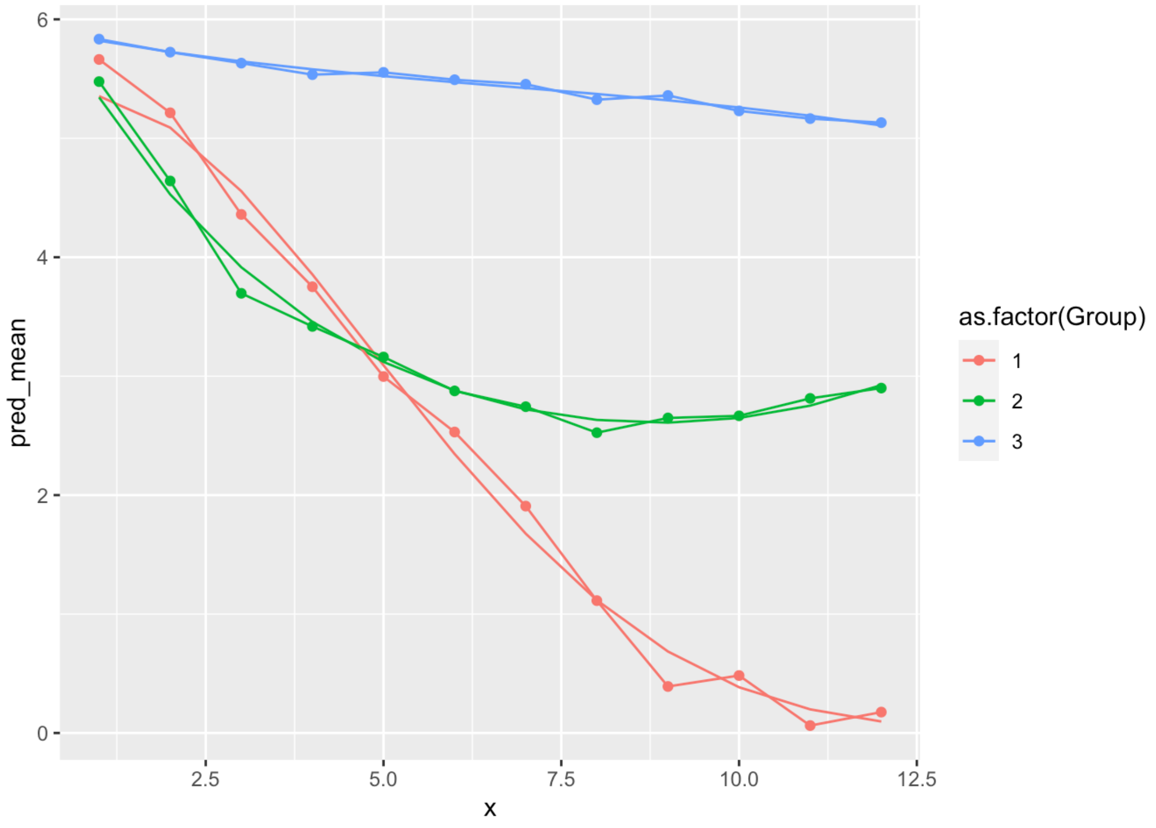


**S F1**, Fit check for 12 week group-based trajectory model. Smooth lines indicate the fitted trend for each trajectory group. Dotted lines indicate the mean value of the raw data at each timepoint within each group.

**S T2**, Coefficients from logistic regression model predicting 6-month treatment outcome based on 12 week adherence pattern, n=596

| | OR (95% CI) | |
|---|---|---|
| | Unadjusted | Adjusted |
| High adherence pattern | Reference | Reference |
| Moderate + Low adherence pattern | **3.78 (2.18, 6.53)** | **3.42 (1.90, 6.12)** |

*The adjusted model included patient sex, resistance category (MDR/pre-XDR/XDR), year of treatment initiation, chest x-ray cavitation (no/yes/missing), sputum grade and BMI. This model did not include region due to positivity violations.*
